# “Shifting and sharing the power” in research: Views and perspectives on research priorities from the Down syndrome, Fragile X syndrome and Williams syndrome communities

**DOI:** 10.64898/2026.08.20.26360980

**Authors:** Laura Cristescu, Elizabeth Pellicano, Jo Van Herwegen, Gaia Scerif, Emily K. Farran

## Abstract

People with intellectual disabilities and their communities are rarely involved in setting priorities for research. Our study addressed this gap through consultations with the UK communities of three genetic syndromes in which intellectual disabilities are common: Down syndrome (DS), Fragile X syndrome (FXS) and Williams syndrome (WS). The study aimed to provide an understanding of (1) the views of the DS, FXS and WS communities on current UK research; (2) their priorities for future research; and (3) participants’ views of engaging with UK research. We conducted focus group discussions with 39 community members including: children and adults with DS, FXS and WS; parent/carers of people with DS, FXS and WS; practitioners and researchers who work with these communities. Our study was carried out in collaboration with a Steering Group and two Advisory Groups of DS, FXS and WS community members. We identified three themes. First, participants shared their dissatisfaction with the current research landscape and wanted a more balanced landscape, with more research with direct application to the daily lives of people with DS, FXS and WS. Second, community members emphasised the importance of translating research into practice, advocating for better access to research and more meaningful participation to research of individuals with lived experience. Third, our study not only identified *what* should be the focus of future research on DS, FXS and WS, but also *how* researchers should conduct their research. Whilst including children in our sample was a strength, there were some limitations to the diversity of our sample; children with FXS were not represented and gender, ethnic and geographic diversity could have been broader. Nevertheless, we hope that our findings will change the future of research in this field so that research carried out in the name of individuals with intellectual disabilities such as DS, FXS and WS, is of direct use to these communities.

## Introduction

There are an estimated 1.5 million people with an intellectual disability^a^ [1, 2] and 3.5 million people with a Rare Disease [3] living in the United Kingdom (UK). Ensuring that people from these communities lead happy, healthy, and prosperous lives is a crucial objective. Yet, the reality is that they all-too-often experience deep inequalities and injustices, including high mortality rates [4], health inequities [5], and high rates of homelessness [6]. Insofar as research is conducted about people in such situations, the stubborn persistence of these inequalities raises questions about its relevance and reach. This is an instance of the problem identified by Nature [7] of “too much research done in the name of society” failing to be of direct use to society.

Participatory approaches are one way of ensuring that research is targeted towards those areas that people with intellectual disabilities and their families identify as meaningful to their lives. These approaches challenge conventional research methods by moving away from conducting research “to” or “for” participants, towards collaborating “with” the communities with lived experience. They draw upon community members’ *practical wisdom* at all stages of the research process, and acknowledge the value of different perspectives in the co-creation of knowledge [8].

Participatory approaches therefore seek to disrupt long-standing power imbalances in knowledge construction by ensuring that communities, including the individuals themselves, their families, and those who support them, have a role in shaping the research that impacts them – including identifying priorities for research. The shift towards participatory approaches is also reflected in the priorities of UK research funding bodies, with many funders publishing guidance to undertake this type of work (e.g., guidelines from the National Institute of Health Research [9]).

In the spirit of participatory approaches, the past decade has witnessed an encouraging increase in research priority-setting exercises, conducted with communities with varying lived experience [10]. A recent review identified 41 such exercises conducted between 2008 and 2020 with representatives of the neurodevelopmental community including intellectual disabilities, autism, attention differences and many more [11]. Of these priority-setting exercises, two were completed in partnership with the intellectual disability community [12, 13] and one with the Down syndrome community [14]. The latter three studies used different methodologies to conduct the priority-setting exercises: an adapted James Lind Alliance (JLA) methodology [13], the Child Health and Nutrition Research Initiative (CHNRI) methodology [12], and an exploratory and descriptive mixed-methods approach [14].

With reference to community members within these three reports, they tackled the existing research power imbalance in research in different ways. While Sinclair, McCullough [14] engaged with parents and Tomlinson, Yasamy [12] with an expert group of professionals in developmental disabilities, the priority setting exercise by the Royal College of Speech and Language Therapists (RCSLT) [13] is the only one to have involved adults with intellectual disabilities themselves, as well as parent/carers and professionals.

The RCSLT [13] study, focused on identifying research priorities relating to communication and swallowing, found that representatives of the intellectual disability community wanted to see research on interventions, services and inclusive communication environments. Similarly, Sinclair, McCullough [14] found that parents of individuals with DS prioritised research with practical application to daily life, such as research that targeted education, dietary supplements, exercise and early intervention. On the other hand, Tomlinson, Yasamy [12] found that professionals working in developmental disabilities, which includes intellectual disability and autism or other pervasive developmental disabilities, identified early detection in low and middle income countries as their top priority for research, followed by research on early interventions and support for families.

Early intervention and service provision, particularly collaborations between service providers, have also been identified as top areas of priority in other JLA Priority Setting Partnerships, such as Childhood Disability [15], Learning Difficulties [16] and Childhood Neurological Conditions [17], although none have been conducted with the intellectual disability community [18]. Furthermore, a recent review of inclusive health and social care research found only 17 studies in which adults with intellectual disabilities undertook research alongside academic researchers since 2000 [19]. Of these, less than half involved people with intellectual disabilities in deciding the research topic, and only two studies involved community members in funding applications [19].

### The current study

Given that so very few priority-setting exercises have been conducted with the intellectual disability community, our study sought to do just that, focusing on three genetic syndromes in which intellectual disabilities are common: Down syndrome (DS: the most common genetic syndrome); Fragile X syndrome (FXS: the most common inherited form of genetic syndrome); and Williams syndrome (WS: a rare genetic syndrome with increasingly early diagnosis). We selected these groups as model syndromes because they have a definitive diagnosis that can be genetically confirmed early on in development and collectively reflect the wide range of ways in which intellectual disability affects life outcomes.

The priority setting exercise itself was one key component of the broader project entitled, *Shape Research, Change Lives: Setting priorities in genetic syndrome research* [20] and, to our knowledge, is the first to explore the research priorities of the DS, FXS and WS communities. The full project involved an initial review of the UK research landscape for the past decade, and documented the profile of research topics in published research and funding awards. We determined that the majority of UK funding and research publications focused on areas of basic science research, which investigated the processes and mechanisms that support the brain, body, and cognitive functions. Research with more direct and practical implications to daily life, such as studies on the development of treatments and interventions, the provision of services, lifespan and societal issues, received little attention.

We then conducted a consultation with members of the DS, FXS and WS communities, via a questionnaire and focus groups, to understand what they thought of the current research landscape and what the priorities should be for future research. The outcomes were published as accessible public-facing reports (see: https://www.surrey.ac.uk/research-projects/shape-research-change-lives-setting-priorities-genetic-syndrome-research#outputs). This approach – including an exhaustive review of the pre-existing research landscape against the research priorities of a range of stakeholders – is what Embracing Complexity and Sapiets [11] described as “gold-standard” for establishing research priorities. Using a “gold-standard” approach means engaging the most appropriate, trustworthy and reliable methods to answer a research question.

In the current study, we report the community members’ views of the current research landscape, and their research priorities from the focus group discussions. We use ‘community members’ when referring to participants across all groups involved in our study. When findings are specific to one participant group (e.g., children with DS/FXS/WS), we explicitly state this. Critically, we built on existing priority-setting exercises by eliciting directly the views of a range of community members, including children and adults with intellectual disability themselves. On account of the emerging nature of participatory approaches to research, we also sought to understand people’s experience of being involved in research. Research has demonstrated that collaborating with communities with lived experience can be beneficial at all stages of the research process [21, 22] and can assist in translational efforts, ensuring that research evidence is produced in formats that are accessible for non-academic stakeholders and can be more effectively applied in practice [23]. Furthermore, it can lead to greater trust of the communities in the research process itself [24]. Asking our communities about their experiences of taking part in research in the current study is crucial if we are to continue to gain insight into how best to refine participatory approaches.

In summary, we sought to address the following research questions: (1) What are the views and perspectives of the DS, FXS, and WS communities on current UK research? (2) What are the priorities for future research of the DS, FXS, and WS communities? And (3) what are their views and experiences of engaging with UK research?

## Method

### Participants

Participants were recruited through the research team’s extensive community networks, as well as those of the Steering Group (see below), via emails, newsletters, and social media. Participants completed an online initial expression of interest form, including basic demographic information (age, gender, genetic syndrome), which enabled us to select a diverse group of participants. Thirty-nine people were involved, including 16 (41%) children and adults with DS, FXS, and WS, 10 (26%) parent/carers of people with DS, FXS, and WS, five (13%) practitioners who work with people with DS, FXS, and WS (e.g., doctors, educators, allied-health practitioners), and eight (20%) researchers working in areas relevant to the DS, FXS, and WS communities.

Most participants (n=29; 70%) identified as ‘female’ and ‘white’ (Table 1). Most children and adults with DS, FXS or WS reported completing primary/secondary or sixth form/college education, while most parent/carers, practitioners, and researchers were well educated. Six children with DS (n=3) and WS (n=3), aged 11-14 years who mainly identified as ‘female’ (n=4; 67%) took part in group-based discussion. Practitioners and researchers had between five to 29 years of experience working with individuals with DS/FXS/WS. Practitioners worked in education, health, and social care settings, whilst researchers worked in the fields of psychology, linguistics, education, health, and psychiatry. Participants lived across the UK, with all four nations being represented and most residing in England (n=32; 82%). The sample is thus not fully representative of the UK genetic syndrome community, with the absence of children with FXS and limited representation of males, individuals from minority ethnic groups and those who reside outside England.

**Table 1.**
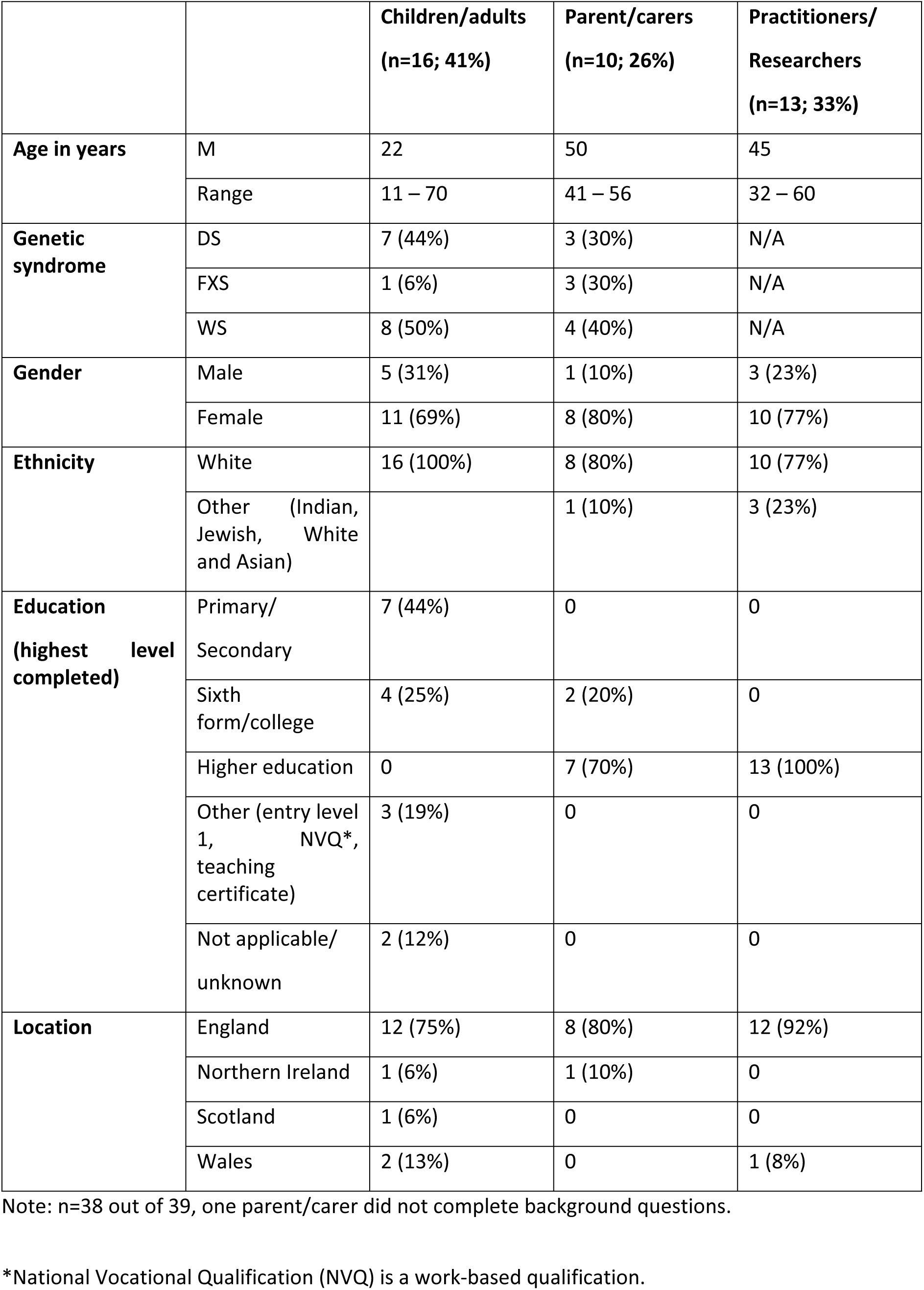
Demographic data for participants that took part in focus groups and interview.

### Procedure

Ethical approval for this study was obtained via the University Ethics Committee (UEC) at the University of Surrey (FHMS 22-23 222 EGA). All participants consented via online consent forms and were provided with a developmentally-appropriate research information sheet. Parent/carers of children and young people under 16 provided parental consent and verbal assent from child participants was obtained during the focus groups. All project materials can be accessed on the Open Science Framework [25].

We conducted 11 focus group discussions, each with three-to-five participants, and one individual semi-structured interview^b^ between November 2023 and March 2024. The focus groups for children, adults and parent/carers were held separately for each genetic syndrome group, while practitioner and researcher discussion groups were mixed, as most had experience of engaging with more than one genetic syndrome group.

The focus groups with parent/carers, practitioners and researchers each followed the same structure. First, we facilitated an initial discussion of their perceptions of research in the field, including their awareness of research, experiences of engaging with research findings, their priorities for research, and the perceived benefits of research participation. Next, we presented the findings from a review of the current landscape of UK research for the three genetic syndromes between 2013 and 2022, including projects funded and publications [20]. Following this presentation, group members discussed their views on the findings and how they align (or not) with their own research priorities. Finally, we discussed the perceived gaps in research and their views on how research and funders should engage with communities with lived experience in a meaningful manner.

We adopted a different structure for the discussions with the children and adults with DS, FXS, and WS. Specifically, ahead of the discussion, we sent parent/carers or the adults themselves a bespoke social story and asked them to use this story to prepare their children/young people or themselves for the discussions. The social story introduced what research is and what researchers do (see [25]). It then introduced the researcher/facilitator, provided an overview of the running of the meeting (including allowing time for breaks), and listed the ice-breaker activity, the open-ended questions (listed below), and one example of a character story (described below). Parent/carers also received the ‘What is a Researcher? [Short version]’ video, along with an online consent form [26]. These materials supported a wider range of children and adults to participate as they reassured parent/carers that the discussions will be carried out at an appropriate level. They also allowed children and adults with DS/FXS/WS to know what to expect from the process, thus reducing anxiety of the unknown.

The discussion sessions first re-introduced the social story slides, and were followed by a discussion of the consent process. Next, we facilitated an ice-breaker activity (e.g., sharing favourite food/colour, singing a song of choice, asking each other a question) to gain rapport among the group. Children and adults were then asked to consider: (a) what do you want to know about DS/FXS/WS? and (b) what do you want other people to know about DS/FXS/WS? Finally, they were asked to rate their research priorities. To facilitate these ratings, we presented eight areas of research through short stories, each told from the perspective of one main character. These areas represented the full range of research topics covered in the review of the research landscape. Participants ranked them using a show of hands: one finger for ‘not important’, one hand for ‘it’s okay’, and two hands for ‘it’s very important’. They were invited to discuss their ranking after each story was told. The characters were revisited at the end, and participants were invited to choose their favourite.

Adults had the choice of attending the group-based discussion independently, or with a trusted adult. For children and young people, parent/carers were required to be present, although it was emphasised that the group-based discussion was seeking the children’s, not the parent/carers’ views. The individual interview followed the same structure. The structure and content of these discussions were shaped collaboratively with the two Advisory Groups (see *Community Involvement* below).

All focus groups and the interview were conducted by the first author, held online via Microsoft Teams. Focus groups ranged from 37 to 85 mins (M = 56 mins) and the individual interview lasted 54 mins. All discussions were recorded with the participants’ prior consent, and subsequently transcribed verbatim.

### Community involvement

The project was carried out in close collaboration with one Steering Group and two Advisory Groups. The Steering Group included members from the DS, FXS, and WS communities with lived experience and/or advocacy and/or policy/funding experience. The Steering Group influenced strategic decisions and supported recruitment and the dissemination of findings. Our two Advisory Groups of experts with lived experience, one with adults with DS, FXS, and WS and one with parents/carers, also met at the same key stages of the research project. The Advisory Groups advised on the methods’ design, participant recruitment, and dissemination. Specifically, their suggestions included the use of social stories to explain the timeline of the in-depth discussions, visual supports with both plain text and images to accompany verbal communication, ice-breaker activities and music to facilitate participants to feel at ease, as well as to allow time for breaks when needed, and to encourage turn-taking. Suggestions were prioritised by the Advisory Group of adults with DS, FXS and WS, based on what they found to be most useful during their meetings. Following their feedback, we created social stories ahead of each meeting that clearly set out expectations and the meeting agenda. A member of the parent/carer Advisory Group commented on the length of the survey and suggested reducing the number of questions included. Implementing this suggestion would have meant reducing the number of research priorities rated and was in contradiction with our research design and aims. To mitigate this, we decided the amend the layout of the survey to reduce scrolling thus reducing the time taken to complete. Feedback from both Advisory Groups was positive following this change. All other suggestions were implemented by the research team both for further Advisory Group meetings, and the participant discussions.

## Data analysis

We analysed the focus groups using reflexive thematic analysis (Braun & Clarke, [27–29]). The analysis was carried out by LC and EP in collaboration, through “deep and prolonged data immersion, thoughtfulness and reflection”, in which researchers’ subjectivity was acknowledged and understood as a resource [29]. We used an inductive approach, without coding to a pre-existing framework, to identify meaning patterns in the data. The data analysis was informed by the authors’ experience and training in education and psychology. Our epistemological stance fits within an essentialist framework, in which we strove to authentically report the meanings and experienced reality of the participants. Our views align with those of the social model and neurodiversity paradigm, through which we recognise the interplay between the individual and their environments, acknowledge neurodiversity and advocate for a strength-based approach. The themes and sub-themes presented below are understood as “creative and interpretive stories”, which are not waiting to be found, but rather created at the intersection of the researchers’ knowledge and theoretical underpinnings, and the data itself [29]. The two researchers double-coded a sample of the data and met regularly to discuss the patterns of shared meaning and understanding. For example, the code “Fighting for a diagnosis” was given to the following quote from an adult with WS: “my mum had to fight, fight, fight 18 months to get me diagnosed with Williams Syndrome…it shouldn’t be such a fight to get everything sorted” (WSA5). This was then included in the subtheme 1.1: “It shouldn’t be such a fight”, along with other codes such as: Annual Health Checks, Worries about the future, Early intervention and More research into access to adult services. EP created the initial thematic framework, which was discussed with LC, and further refined upon discussions with the rest of the research team (JVH, GS and EF). The research team reflected on how own assumptions and experiences might influence the interpretation of findings. The thematic framework was discussed with the Advisory Groups and the Steering Group during meetings, and it gave opportunities for the themes to be further refined. Analysis was therefore iterative and reflexive.

## Results

We identified four themes (Fig1), which we describe in turn below. Throughout, quotes are attributed via ID numbers and the respective genetic syndromes (DS, FXS and WS) and groups (‘A’ refers to adults with the genetic condition, ‘C’ for children/young people with the genetic condition, ‘P’ for parents, ‘PR’ for professionals and ‘R’ for researchers).

**Fig 1.**
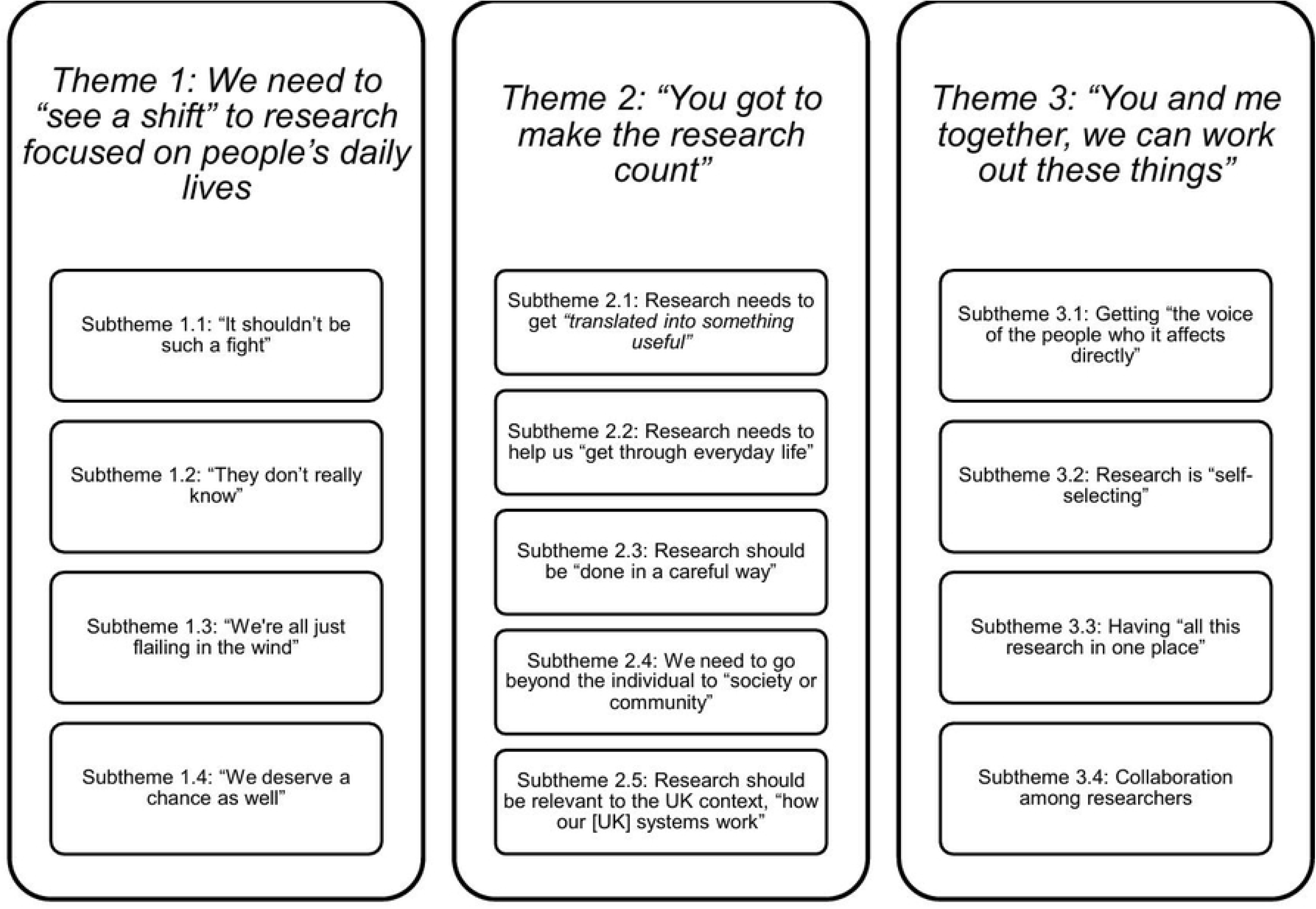
The views and perspectives of the DS, FXS and WS communities on current and future UK research: themes and sub-themes.

### Theme 1: We need to “see a shift” to research focused on people’s daily lives

Community members recognised that, traditionally, research has been focused on biology, genetics and medical research. While they noted these areas represented a “right and proper focus” (PR1), they wanted “to see a shift so that the focus isn’t quite so heavily weighted” (PR1) towards these areas.

Subtheme 1.1: “It shouldn’t be such a fight”

Community members described their constant battles navigating systems and services and the toll such battles caused.

### Fighting for services

These sentiments were rooted in what people described as often deeply challenging lives. Parents and individuals themselves repeatedly reported being “failed miserably” (WSA5) by medical professionals, and education and social care services, which they resoundingly felt “are the barriers to [child’s] life outcomes” (DSP2). They spoke of how they felt they were “constantly fighting” (DSP2) for services. For some, this “battle” (FXSA) began as they tried to access diagnostic services, when “no-one seems interested” (FXSP1), and where “you push and you push” (WSP1) until one professional “finally listens” (WSP1). One WS adult implored, “my mum had to fight, fight, fight 18 months to get me diagnosed with Williams Syndrome…it shouldn’t be such a fight to get everything sorted” (WSA5). The “constant battles” (DSP2) continued post diagnosis. They described how there was “a postcode lottery” (WSP1) “a massive disparity across the country” (PR1) in terms of support. Services in local areas were severely limited or, even worse, absent altogether: “there is nothing for [young adults] in [local authority] (WSP2). One parent explained: “there are people who say, ‘early intervention is key’, you’ve got to be doing this, you’ve got to be doing that, and then I would go and hit a blank wall with the services within my area” (FXSP1). Similar issues were experienced for healthcare: “GP practices now by law have to offer an annual check-up to young people with learning disabilities…but it doesn’t always work because not every GP practice is cognisant of this” (WSP4).

### A lonely fight

Parents also emphasised how the weight of responsibility to secure services for their children with DS, FXS and WS all-too-often “lies on the parent” (WSP4): “we got left with the diagnosis and had to find the rest from breadcrumbs left along the way” (FXSP2). What they wanted was “people who know what they’re doing and to say, ‘this is what you need, you need this and this, and you need to do this’ – just a bit of a support on your shoulder” (DSP1). In the absence of such support, the fighting sometimes continued until parents “haven’t got anything left to give” (DSP3). One parent went so far as to say that “this is what [local authorities] prey on…the parental exhaustion” (WSP4). Of the few parents who reported receiving either an early diagnosis for their child or the services they felt their child needed, they reported feeling “very fortunate” (WSP3) and “lucky that somebody finally listened to us” (WSP1).

### Fighting for the future

The situation was felt to be particularly dire for young people transitioning to adult services: “at 18, you’re just left in limbo” (WSP1). As one parent put it: “it’s a bit of a black hole” (WSP1). One parent lamented how her child and “a lot of his friends have been through an awful lot, just trying to access supported living, independent living, but there’s no research going on about how that affects them as a human being” (DSP2). WS respondents also noted that the lack of adult services affected their access to community life, including how “our public transport is a shambles” (WSA5), particularly in regional areas, where they wanted transport run by “people that understand people with disabilities” (WSA3). Given the enormity of these challenges, researchers and professionals felt that community members wanted research on “what’s coming next for them, or what’s causing them barriers, which is why transport is such a big deal, and housing” (RS5) and the need for “systems-based research on how we support and develop services for those who can’t access them in the same way that you and I do” (PR1).

### Subtheme 1.2: “They don’t really know”

Having to fight for services was compounded by their encounters with professionals who our interviewees perceived to lack knowledge about their or their children’s conditions, highlighting a need for training and overcoming misconceptions.

Community members felt this was a significant barrier to people with the three genetic syndromes receiving the right support at the right time. One parent stated: “I would say in our daily lives, about 30% of our battle is dealing with the issues that surround [child] with his additional needs, and the other 70% is probably battling against professionals who are either disinterested or lack knowledge entirely” (FXSP2).

### Lack of understanding of co-occurring conditions

Parents were frustrated by how “they [professionals] don’t really know” (FSXP1): “How is it that you’ve got a paediatrician that doesn’t even know what the pathway looks like? It’s very scary” (FXSP3), and who were not open to listening: “it’s like banging your head against the wall…to get people to listen and then flag up what might be going on” (WSP2).

The perceived lack of understanding by professionals was felt to hinder the identification of possible co-occurring conditions in children with the three genetic syndromes (“the physical difficulties that someone like my daughter faces on a daily basis don’t always correspond to the manifestation of Williams syndrome”; WSP4) and meant that key health issues could be overlooked: “a lot of the time you get, ‘oh, that’s just because they have Down syndrome’ – and in a lot of cases, it’s not gone any further, and obviously that can have some real serious health issues” (DSP3).

### Moving away from unhelpful assumptions

Stereotypes and misperceptions were also felt to be potentially harmful: “people like me with Williams syndrome can come across as more able than they actually are…and they can misjudge…so I think more understanding to not judge somebody quickly” (WSA4). These misperceptions could also prevent tailored support:

> Often people associate Fragile X as being very autism-like, but it does have differences [which] really come into play with things like teaching strategies…so it’s making sure the people know and are aware – that’s something that can be really valuable to help a child. (FXSP3)

Consequently, community members spoke of how professionals “need so much more education” (WSA5) about genetic syndromes. Overall, they did not expect professionals “to become experts on the three genetic syndromes” (PR1), but “for professionals to understand that there might be something they don’t know about, and it needs looking up” (FXSA). Community members also wanted professionals to “treat them as individuals and work with them at their level” (WSP1). As one researcher put it, “What difference does it make knowing their diagnosis?…it’s more useful to know what their communication styles and preferences are and, you know, their support needs and things like that” (RS7).

### Improving public awareness

This awareness-raising needed to be “available not just to professionals but to people out there in the mainstream population” (PR3), “and into industries, or getting out into the community with, like, supermarkets – how can we support wider community aspects so that they then know how to support anyone that they then come into contact with?” (PR5). Young people agreed: “it’s best that they get it out that [WS] is a thing and stuff – we all need to learn about that” (WSC1). One professional suggested, “maybe we need to do similar work that the UK did around dementia, breaking the barriers and the myths so people’s awareness increased, and they were more friendly and more supportive of families with a loved one with dementia” (PR3).

### Subtheme 1.3: “We’re all just flailing in the wind”

Worries about the future, adult life and old age were commonly expressed. Community members were concerned that the lack of knowledge about DS, FXS and WS meant that they could not be sure of what the future holds for their children or their family: “to be honest, the thing that keeps me awake at night, is what the future looks like, what the roadmap is” (FXSP2).

### No roadmap for the future

They reported being surprised that lifespan issues were not “the driving force of the research” (PR5), and the apparent lack of “research into what really matters for them as young adults” (DSP2) or “into the ageing process” (FXSA). This lack of research “to do with life” (FSXP2) meant that they simply “did not know what’s next” (WSP2) or “what a positive outcome looks like for our young people. You know, what are people happy with, what’s reasonable, what’s active, what’s fulfilled, what’s healthy, what’s supported? What are the positive role models we can draw from?” (WSP1). They reported feeling desperate “to have somebody just say this is what could happen in the future…this is what’s going to happen between this age and that age. Just helpful hints” (WSP1). Professionals agreed that families needed to know “what they can do now they’ve got the condition, what they can do about moving forward and making sure that they can be included and live their life as prolonged as possible” (PR5).

### Fear of the unknown

In the absence of this knowledge, parents were therefore anxious about “making sure that [child’s] future is secure” (FXSP1): “it’s that whole, are we doing right? You just don’t know. You do the best you know you can do given the information you have” (DSP1). For many, this uncertainty led to “fear”, and even to parents blaming themselves: “because there’s so much unknown, maybe you’re the one that’s doing the bad thing. Maybe you’ve not done enough, and that’s why your child’s not speaking” (FXSP1). Another parent said:

It’s like a jigsaw, isn’t it? We’re all making up a jigsaw and we don’t know what the pieces are, what we’re trying to build, but we just want to get somewhere and not look back at our lives and think, well, we could have done a better job of that. (WSP2)

### Impact on siblings

Parents were also worried about the effect this uncertainty had on their family’s wellbeing, including for siblings and the “emotional weight” (FSXP3) they may feel about what the future holds for them: “I worry about is that [sibling] will be the one left looking after everyone after we’re gone. And I think that already scares him. It scares the hell out of me” (FXSP1). One parent, however, disagreed: “you know, people are like, ‘I feel so sorry for the siblings’, but do we need to feel sorry for the siblings? Are they actually worse off? Or are they, in fact, better off? Because what I’m seeing is lovely” (DSP1).

### Subtheme 1.4: “We deserve a chance as well”

Despite this uncertainty, community members highlighted that people with DS, FXS or WS “do have a part to play in society” (WSP2) and wanted to see research in areas that would allow them to “live life to the fullest every day because we don’t know exactly how long we live for” (WSA1).

In the words of one WS child: “everyone should have a role in their life…We deserve a right to get a job. We deserve a right to have a roof over our heads and get paid…We deserve a chance as well” (WSC1).

### Seeing the person, not the stereotype

Instead, children and adults with DS, FXS, and WS were concerned that these aspirations were undermined by pervasive stigma and bullying: “people mock people with learning disabilities. It doesn’t matter if it’s Williams Syndrome, or different intellectual disabilities, it’s not funny. Having a disability and having learning issues is nothing to laugh about” (WSA1). They discussed ways of how research could help mitigate these negative attitudes and actions, including increased public awareness so that “everyone knows about Williams syndrome. I want the whole world to know” (WSC1), especially so that it can “help people who are doing their things to help people [with DS]” (DSC1). They also spoke of awareness-raising efforts to combat stereotypes, including about how there is “so much generic around in Williams syndrome with the happy syndrome” (WSA5): “we’re not happy all the time. We do have our struggles. I feel like it’s important people to know that” (WSA3). They also called for more visibility in society, with “more people like on magazines and more models and more fashion showing people have Williams Syndrome” (WSC1), and greater understanding, for “people to be kind, to be patient” (DSA2) and “to talk to [people with DS], about how they feel” (DSC1).

### Difference is beautiful

They also showed pride in their identity, as one DS adult put it: “it’s okay to have Down syndrome” (DSA3). Another recounted how this pride was hard-won: “this year is first year I really fully enjoyed having WS, it took me so long to love it really” (WSA5). They emphasised the importance of valuing difference – “no matter how different you are, you’re always gonna be yourself and you’ll be beautiful and don’t feel like you’re ugly because you’re not ugly” (WSC1) – and seeing the person, beyond the label: “we are not labels, we do not need labels to tell people who we are” (WSA5): “Down Syndrome is a small part of me, yeah, I am so much more…it does not define me who I am” (DSA4). Children with DS added that they wanted others to know they are the “same” (DSC2) and can “be friends” (DSC3), and although sometimes “might need some help with some words…words tend to get tricky sometimes” (DSC1).

### Seeing the strengths

During discussions, parents also emphasised their children’s strengths, which they implied was not reflected enough in current research. They spoke of their “character and great sense of humour” (FXSP3) and how “they do just know how to bring on the charm” (WSP2). People also felt that “they’ve got a lot to offer society and they can teach you things you didn’t know you needed to know” (WSP2), and felt they were all-too-often underestimated: “there is so much that they are capable of – sadly, too many assume they’re not. [FXS sibling], in his 60s, learnt to tell the time. And he can still do that” (FXSA).

### Theme 2: “You got to make the research count”

Subtheme 2.1: Research needs to get *“translated into something useful”*

Our interviewees’ stark descriptions of the everyday challenges they faced set the context for their views on research, namely, that they wanted research to make a meaningful difference to their lives.

### Research as a valuable tool

Parents felt that taking part in research is “valuable” and “useful” (FXSP3), helping them “personally” with understanding of development (WSP3) and “psychologically [to] feel like we’re being proactive” (FXSP2). They also felt research helped them “practically” (FXSP2), by enabling them “to find evidence” (FXSP1) to help with accessing services, when “going to tribunal to try to get him into a school that has other children with FXS” (FXSP1), and with “knowing what to put in the Education, Health and Care Plan” (WSP1) or “Disability Living Allowance applications” (FXSP3). In this way, they felt that relationships with researchers were beneficial – “ammunition of the backup of a professor studying [FXS]” – and a source of “help from the professionals who are interested in the things that affect us” (FXSP2).

### Moving research findings from paper to daily life

Community members also emphasised that more work needs to be done to ensure that research “moves from being on paper to actually getting out there and helping the community” (FXSP3). It was recognised that this can be a “massive challenge” as “there’s so much valuable data, that it’s just getting it out there to the right people” (FXSP3). Instead, it is essential that the focus shifts “to the practical and the applicable” (PR1) to provide individuals with genetic syndromes with the opportunities to “live their best life or their life to the best of their fulfilment” (PR1). Some researchers admitted that their work is “unpractical”, or they are “at least a step away from anything really practical and important” (RS1). Parents agreed, and whilst recognising the value of genetic research and how “it’s exciting when you see that happening…it feels very distant, it’s in the trees” (FXSP3). One community member summed it up: “there seems to be too much focus on the why, and not so much the how” (FXSP2).

### Disseminating research to professionals

Community members recognised that the challenges in translating and implementing research findings “into something useful” are in part because they felt there is currently “no bridge between the academic research to the actual community professionals” (FXSP1). Moreover, they felt that extensive efforts should be made to share the information “within the professional community effectively and quickly…so that there is a pot of knowledge for specific conditions like this” (FXSP2). Although they recognised that some areas of research, such as cognitive and genetic research have received significant attention, community members stressed that “nobody’s really taking any notice of it” (DSP3), with some recommendations “totally ignored by practitioners” (DSP3).

### Subtheme 2.2: Research needs to help us “get through everyday life”

Community members agreed that research should be aimed at supporting people to “get through everyday life” (WSA1), focusing on access to adulthood opportunities, and appropriate mental health and wellbeing support.

### Improving adulthood opportunities

The importance of research and provision of services to support with the development of “independence” (PR5), “daily living” (WSP2), “preparation for adulthood” (PR5) and “life skills” (DSP2) was discussed at length by community members. It was seen as essential in ensuring good “quality of life” (PR4), enabling people with genetic syndromes to “have better lives and do things that other people can do” (PR2), to “access the same opportunities” (PR5) and have “all the right tools to reach [their] full potential” (DSP3). Parents added that practitioners have shared information on “this happens because of this syndrome…but nothing to do with how you go about daily life” (FXSP1). Adults with genetic syndromes emphasised that “being independent is important” (DSA1), some sharing their difficulties with accessing housing services (“I’m trying to get into accommodation and live by myself at the minute”; WSA5), and others their success stories: “I live by myself, I am an independent person with WS” (WSA1). Adults were resoundingly clear that people, regardless of their genetic syndrome, shared aspirations about “working a job” (DSA2), managing “my own money” (DSA1), and “living in a flat” (DSA3) or “with my friends” (DSA1). Children with DS and WS shared similar aspirations, viewing having a job or living independently as “very important” (DSC3) because “everyone in life can have a job” (WSC1).

### Better understanding of wellbeing and mental health

“General wellbeing” (PR4) and mental health (WSA4), particularly anxiety and depression, were agreed to be “high topics” (WSA5) for research. One child with WS wanted to know if anxiety “gets worse or better as you get older…because I have lots and lots of it” (WSC5), and another was seeking an answer for: “why am I anxious? That’s what I ask myself every day” (WSC2). Parents agreed and wanted to understand “the different types of anxiety” (FXSP2), “what triggers their anxiety…and looking at ways to help manage it” (FXSP3). There was a call for more understanding of “the link between mental health and WS” (WSA4). One child with WS summarised the value of research on mental health as: “Very important to me as well because I get very anxious and I hate the overlapping feeling, like it makes me feel like I want to be sick, like, that’s how anxious I get, and that scares me” (WSC1).

They also shared their worries about “struggling with emotions” or “getting angry really easily” (WSC1) and wanted research to help them understand these issues. Participants highlighted the importance of more awareness and understanding in society, for people to “not stare at us when we get angry because it can get very annoying” (WSC2). They highlighted the importance of having “friends and the amount of support that I need to help reduce [my anxiety]” (WSC1) and having “proper normal adult friends” (WSA5). Practitioners and researchers added their interest in “designing an anxiety intervention” (PR2) and called for further understanding of the “presentation of different mental health conditions in these [genetic] syndromes” (RS7), and the “parity of esteem between mental health services and physical healthcare services” (PR1).

### Subtheme 2.3: Research should be “done in a careful way”

Community members were “surprised” (RS2) at the lack of research on treatments and interventions, which community members “haven’t seen much on” (WSP2) and “thought it would be a bit more” (DSP1). They highlighted the importance of a person-centred approach, and the understanding of the individual’s unique characteristics “to know how to tackle things well” (WSP2).

### A call for appropriate interventions

Parents wanted to know if there was a “better way of doing [daily living tasks] with someone with very poor fine motor [skills]…or coming up with a crafty way of building it into your life” (WSP2). There was a call for more understanding of related health conditions and characteristics, such as “dysphagia…posture and gait issues” (WSP2), “musculature issues” (WSP4), “spina bifida, scoliosis and kyphosis” (WSP1), and the tailoring of suitable interventions. Although there was agreement on the importance of daily living skills, community members acknowledged the differing wants and needs of people with genetic syndromes. One parent shared that their teenager was “focusing on daily living and life skills and actually working, but he’s not ready for that” (WSP2), whilst another recounted that their young person has “certainly outdone his life skills. He did it in specialist school. He did it at college twice. It was the only course that was available…it’s like how many life skills can this young man be taught?” (DSP2).

Adults with WS also advocated for awareness and understanding of individual strengths and needs in accessing opportunities, which has “to be done in a careful way, because it’s important that people don’t jump the gun with seeing how able the person is” (WSP4):

> You can’t just tell a person with WS: right, you need to go and live by yourself, you need to show other people competence. It does not work that way at all. It’s the way of sitting down with someone and actually taking some time to actually talk to them on a one-to-one basis, every one to two weeks and trying to explain to them what’s going to happen. (WSA1)

### Development of understanding and person-centred approaches

Focusing on a person-centred approach is also relevant to accessing suitable services such as “school setting” (FXSP3). Practitioners specified the value of teaching behaviours that promote independence, as young people can “get so used to working with a teaching assistant one to one” (PR4) and that of friendships with peers: “it’s nice when you got new children starting school, they suddenly got a new friend, and they can walk down the path together holding hands” (PR5). One parent/practitioner shared the lack of understanding and conflicting evidence she found regarding children with FXS learning to read phonetically or through “sight reading”, and the value of “teaching them slightly differently, then maybe they can [learn to read]” (FXSP3). The person-centred approach should also be applied to adult services, as one parent/practitioner noted: “the kids I’ve been working with as they’re getting older, they’re getting less fluent, they’re developing stammers and stutters” (DSP1). One adult with WS summed up the range of health needs: “some of us live for a very long time, some of us go through a lot just to stay alive” (WSA1). Other lifelong areas of priority for research discussed included “long term memory and communication strategies” (PR5), “language” (RS1), “benefits and housing” (RS5), “sexual activity” (PR4) and “support to have a family” (RS8). Overall, community members agreed on the need for providing the “same opportunities” (PR5), such as “prolonging education” throughout all life stages (FXSA) to enable “the environment to be more accepting and tolerant and to be more inclusive” (RS4).

### Subtheme 2.4: We need to go beyond the individual to “society or community”

In keeping with community members’ rich descriptions of the everyday challenges they faced (Theme 1), they emphasised that research should not just focus on “figuring out what’s wrong with me” (WSA5). Instead, they wanted research on DS, FXS and WS to focus on understanding the different contexts in which people grow up, study, work and live. They felt that this more holistic view – understanding the individual and “society or community” (DSP2) – would help identify the environments that match a person’s needs, preferences and interests:

> I’d be interested to find out what people with Down syndrome think about what Down syndrome means to them, self-identity and how they feel that they’ve had to fit into society. I think that would be a real turning point for services. (DSP3)

### Understanding the role of the environment

Community members noted that research should help with understanding how people with genetic syndromes “bring people together…[they] do have a part to play in society and they do make people feel good about themselves…There’s a magic in there” (WSP2). Understanding the characteristics of the environment that contribute to offering opportunities for flourishing is essential because: “in the right surroundings the Williams person is great; they are really sociable” (WSP2). This goes hand-in-hand with the person-centred approach discussed above by recognising the interaction between the unique individual and their environment, and moving away from “diagnostic overshadowing” (DSP3). Some community members recognised that “the way society has changed since we grew up, it’s amazing and more of the positive” (FXSA), whilst others stressed the difference that increased awareness could make:

> I can’t help but wonder if that would be the case if society thought differently, you know, if everybody just saw, not just simply as a difference, of course, it’s complicated to raise a child with a disability, but if there was much more support, if it wasn’t seen so negatively, would that still be the case? I think that’s the big question. (RS2)

### Subtheme 2.5: Research should be relevant to the UK context, “how our [UK] systems work”

Community members emphasised the importance of conducting research in context, to support translation on findings to UK practice and systems.

In emphasising the environments in which people are embedded, community members made comparisons with service provision in the United States (US), which left parent/carers wondering how their child’s development might look in a different context:

> In the US, some children you hear about having like four hours a day of therapy, and it’s like, wow, that’s a whole different world. I wonder how that the like the outcome for that child differs to the outcome from here in the UK, where we get speech and language once every six months, and they just observe the child and go, yeah, he’s still not speaking, is he? (FXSP3)

### Conducting context-appropriate research

They noted how other countries, especially the US, “do stuff differently” (DSP1) and “they tend to approach issues quite early on” (FXSP3), which meant that services, including early intervention, are “never going to translate to over here” (DSP1). They emphasised that research should be appropriate for the UK context and “how our systems work” (WSP2). Parents were aware that, in the US, the “families are very instantly put in for speech and language, occupational therapy…as soon as there’s a diagnosis” (FXSP3) and that there was a greater focus on basic science research, with “loads of money going into mouse models of things like DS” (RS1), and greater opportunities for medical interventions:

> We would love to have been able to create a study for low dose sertraline to help anxiety…it’s not like we’re desperately wanting to give our children drugs, but there is research already out there in the US where it can help language developments.

And obviously, having spoken to quite a few of the doctors and the professionals out in the US now, they have all amazed that we have never been had the opportunity to try sertraline for [our son] to help with his speech development. He is now six and he is still nonspeaking; it’s starting to feel like it is potentially a bit of a missed opportunity. (FXP3)

### Theme 3: “You and me together, we can work out these things”

Our community members had strong views not only on what they wanted research to focus on but, critically, also *how* the research itself should be done. One child with DS summed this up: “you and me together we can work out these things” (DSC1).

Subtheme 3.1: Getting “the voice of the people who it affects directly”

All community members agreed that research should be informed by lived experience: “it’s always important to get the voice of the people who it affects directly” (PR5), whilst also recognising a need for developing infrastructures to support collaborations.

### Beginning to shift the power

Parents described community engagement in research as “potluck” (WSP2), adding they “very much doubt anyone’s asked people with DS [about opinions on research]” (DSP3), with one going as far as saying, with reference to the current study: “you are the first, you are the pioneer” (FXSP1). Researchers recognised that research had previously been “driven by researchers and what they wanted to research…but that is starting to change” (RS7). They emphasised that community input in research – when done in “a meaningful way” (RS5) – is “crucial…there’s no point designing a project that nobody wants to engage with” (RS4).

Researchers also acknowledged that more understanding is needed of “shifting that power from what we want to do to actually what might people who have these genetic disorders want us to research” (PR2), as well as “how do we work together and share power in a research project” (RS5). They emphasised that collaborations with communities should take place “right from the start, from writing the grant application” (RS8) and in that way “they might be more invested, and it’s going to hopefully align and meet their needs a bit better” (RS7).

### Improving infrastructures for community involvement

Despite this enthusiasm from researchers, they also expressed concerns they “don’t get enough support…and need more training” (RS3) and “time” (RS2), as well as a supportive “infrastructure” (RS5). Researchers were aware of funders encouraging community consultations, but were uncertain “whether they [funders] do it themselves” (RS4) or “if they do, it’s fairly tokenistic because they [funders] have their own priorities” (RS7). Researchers also felt that since funders are “gatekeepers to the money” (RS7), they should involve the communities earlier in the research process and allocate appropriate funding for doing so:

> Unless [funders] drive [community involvement], we’re never going to be able to pay for our time because so many of the groups that actually consult with people with these conditions are done on a shoestring or by the goodwill of an individual that if they leave it goes, you know, they’re the one that’s building the links to people and the trust. And so yeah, it does rely mostly on people giving their free time and energy. (RS5)

### Collaborations should be meaningful

The lack of resources and understanding of working with community members meant that researchers tend to work with “the same people, the ones that are taking part in everything” (RS5). They compared this shift with that occurring within the autism community, and how autistic people, mostly without an intellectual disability, have become increasingly “visible” (RS2) in research, and how this differs from the genetic syndrome community as most find it difficult “to advocate for oneself and rely on others to advocate for [them]” (RS2). Indeed, researchers emphasised the challenges of meaningful involvement for people with genetic syndromes in research, as “parents still take a very prominent role…making those decisions themselves” (RS3).

### Subtheme 3.2: Research is “self-selecting”

Community members spoke, too, about the lack of diversity in those participating in research. They explained how opportunities to take part in research are often accessed by a small group of individuals who are motivated to contribute to change and have the means to do so:

> There are pockets. Even within research, it’s self-selecting in itself, because you’re going to get those parents that want to make a difference, the ones that know what a difference will make. You’re not going to the ones, the opinions that you really should have. (DSP3)

### Broadening community voices

Community members wanted future research to be “more inclusive” (PR3) of everyone, including “the ones that don’t speak out, or don’t know how, or don’t” (RS1) and “people with more moderate and profound learning disabilities” (PR3). Practitioners and researchers acknowledged the barriers to doing so, including providing accessible information about research, “capacity to consent” (PR3), “inclusive research practices” (RS7) and methods, such as using “visual data pictures” (RS6) and measuring neural activity. Community members felt that research is often “not accessible for the people who the research has been done about…So just thinking about different research methodologies, and broadening that out a little bit, it’s something that I’d like to see” (PR2).

### Subtheme 3.3: Having “all this research in one place”

Community members expressed “a very limited understanding of the scope of the research” (PR1) and a perception that “there’s not a lot going on…there’s just not a lot of information out there” (DSP2). They emphasised the importance of making research findings accessible and centralised.

Information regarding research was often accessed via newsletters from charities and local groups, by “seeking out information” (PR5) on a topic of interest, from previous research participation (DSP2), often organically depending on the “people who are around you” (PR2). One parent commented that: “so while I think the research that’s taking place is informative, I just feel the way that it’s been then given to parents or organizations isn’t timed very well” (WSP1).

There were also calls to make information about research accessible and centralised and to improve visibility and transparency, and “when information is found, to spread it within the professional community effectively and quickly” (FXSP2). Community members lamented that they have “to go out and look for research myself” (PR3) and that doing “all your own research…is an awful lot of effort on parents” (DP2). While they acknowledged that research is often necessary to access appropriate services (see subtheme 2.1), many shared their struggles on knowing where to start because “you don’t know what you don’t know” (WSP3). Parents spoke of their frustrations with having to “find evidence to show what would be beneficial” (FXSP3) and wanting to “actually know what each parent has and gets for their child” (WSP1).

### Subtheme 3.4: Collaboration among researchers

Finally, researchers discussed of the value of collaboration among researchers and professionals from different fields.

Children and adults with genetic syndromes shared it is “very important” that “many people work together on DS/FXS/WS research”. Some chose this as their top priority: “it was the best because we need everyone to be included and learn about DS” (DSC1), whereas others were more sceptical: “I don’t think it’s gonna happen” (WSA5).

Researchers explained that “multidisciplinarity is something that we’re not kind of doing right, we’re not seeing enough of” (RS1), and this represents a “gap” (RS4) in genetic syndrome research. They felt this type of collaborative research that includes different perspectives is critical to making progress in the field and is “really important” (RS4), but is “the hardest to actually get funding for” (RS2).

## Discussion

Overall, our focus group discussions with members of the DS, FXS, and WS communities clearly showed they were dissatisfied with the current UK research profile. Participants expressed frustration at the volume of research on basic science alongside surprise at how little research has focused on treatments and interventions. As a result, they wanted to “see a shift” for more research focused on areas that would have a more direct impact on their everyday lives, thus advocating for a more balanced research portfolio. While they agreed that both basic science and applied research were essential and mutually-informing, they nevertheless advocated for a more balanced research portfolio. Basic science research is an essential building block in areas of applied research, such as developing interventions. The two are not in competition, but rather complementary and can be mutually informing. Community members also wanted research to be more effectively translated into policy and practice, with real impacts on UK systems and processes, and emphasised that, for these changes to happen, researchers needed to work together *with* community members – including children and adults with DS, FXS and WS themselves – to set the agenda for what gets researched and how that research gets done.

### Shifting understandings of DS, FXS and WS research

Our participants repeatedly spoke of the need to address a range of crucial social issues across several different spheres, including healthcare, employment, housing, transport, as well as more individual issues (although, notably, some went further to suggest there was too much research focused on “figuring out what’s wrong with me”). This perceived lack of emphasis on broader issues of this kind could be a consequence of the approach typically taken in scientific research on DS, FXS and WS communities. Such research has predominantly been conducted within a conventional medical model, in which the behavioural, cognitive and neural functioning of individuals with DS, FXS or WS are frequently compared to some typical or ‘normal’ level of ability that is held as the ideal ‘state of health’. Under this model, interventions and treatments are typically designed to remediate these apparent shortcomings and bring people’s functioning in line with the accepted norm. One problem with this approach is that it places undue emphasis on the specific attributes of DS, FXS and WS individuals as opposed to the broader contexts in which they actually live – precisely as our participants attested. Indeed, the everyday challenges they so richly described (Theme 1) were felt to be just as frequently consequences of the way that the world is set up as they are the direct result of any personal ‘failing’.

It is not surprising, then, that our participants were frustrated with the current profile of research, which is dominated by biomedical research – that is, research on the underlying biology and causes of DS, FXS and WS [20]. While our participants very much valued such research, they also wanted to see research targeted towards areas that would make a more immediate, practical impact on their everyday lives – on treatments and interventions, especially those targeting life skills and good mental health; on accessible and effective services, particularly those supporting young people transitioning into adulthood; and on cultivating greater knowledge of, and more positive attitudes toward, people with intellectual disability in professionals and in society more broadly. Put another way, they wanted research that not only focused on individuals with DS, FXS and WS, but also on the broader contexts in which they are embedded, especially on the structures within institutions – schools, GP surgeries, workplaces – that might often prevent them from receiving the most effective support. Therefore, the priorities shared by the DS, FXS and WS communities suggest that a ‘shift’ is needed not only in the topics researched, but also in how research is conceptualised – moving away from the medical model towards one that recognises the interaction between the individual and their immediate contexts.

### Communities’ top priorities for research

#### Adulthood opportunities

Our in-depth discussions showed that people with DS, FXS and WS want to be able to participate in their communities – to gain and hold down a job and to live on their own – and to develop the everyday life skills that will enable them to reach these goals. The associated benefits they identified, such as increased independence, pride and having a chance to contribute to society align with those previously identified in the intellectual disability literature [30–32]. Participants also shared their wishes for fair access to appropriate supports and services, to encounter professionals who understand their individual strengths and needs and offer personalised care, and to enjoy their lives free from prejudice and stigma. Unfortunately, our findings also revealed how far people with DS, FXS and WS and their families are from achieving these goals. Community members repeatedly spoke of the significant challenges they, their children, or those they supported faced in accessing services, from diagnostic services and educational provision to the provision of adult services. These findings confirm so much of what is already known: that people with intellectual disabilities have poorer health outcomes, higher rates of health inequalities [2, 5, 33–35], are over-represented in homeless populations [6] and all-too-regularly experience discrimination and delays in care or treatment [35–37]. The challenge in gaining appropriate services was compounded by a lack of awareness and understanding of genetic syndromes and their associated conditions from professionals, which adds to existing evidence of this gap [38, 39].

Future research endeavours should therefore aim to identify and address the above systemic barriers to service access, employment and housing for the intellectual disability community, as well as to develop training and awareness for public service practitioners and the broader public. Such research efforts should be developed alongside community members, drawing from their lived experience to inform the development of resources that can be effectively translated into practice. Most importantly, this work should ensure that individuals with an intellectual disability are at the ‘centre of the conversation’, through processes that support agency, self-determination, expressions of preferences and choices. Researchers should strive for diversity of representation to understand the lived experiences of individuals in different life circumstances and socio-cultural contexts, along with moving away from a medical model construction of research. In the long-term this could help address health inequalities, reduce delays in care and treatment, and the high rates of homelessness.

#### Personalised support

The need for personalisation of support also came across from all discussions related to interventions and services. Community representatives called for a recognition of an individual’s unique strengths, needs and desires, and for this to be central to all support accessed. The strengths of people with genetic syndromes were highlighted, having the power to bring people together and a positive influence on those around them. There was also a recognition of the impact of the environment and a call for more research to understand its role in development, moving away from a focus on any individual ‘failing’ and medical model research. This extended not only to the individual’s immediate environments, but also to the wider UK systems and a better understanding of their role in the provision of interventions and services. Despite UK policy supporting the personalisation of social care and support services, in practice there is much progress yet to be made [40, 41]. Funders and researchers should focus their future efforts on better understanding the challenges to providing personalised support. Intellectual disabilities are common in the DS, FXS and WS communities; the development of interventions and services for these groups could therefore potentially benefit the wider intellectual disability community.

#### Mental health

Research on understanding mental health, and the provision and access of appropriate services was another top area of priority. This adds to existing evidence of the high prevalence of mental health conditions in the intellectual disability community [42, 43]. Dealing with anxiety, depression and managing emotions were common challenges experienced by the community, which impacted the quality of their day-to-day lives. Drawing on Theme 2, there was a unanimous call for more research to understand the unique presentation of mental health conditions in individuals with genetic syndromes, as well as improved access to personalised services, and awareness from professionals and the general public. This is in accordance with prior research which identified a lack of service availability [44], and a limited understanding from professionals [45, 46]. Mencap also documented a lack of accessible communications or other reasonable adjustments as limitations to accessing health care services and personal support for people with intellectual disabilities [1]. This is despite government guidance on reasonable adjustments for people with an intellectual disability [47] and the commitment of the Equality Act 2010 “to increase equality of opportunity”. Future research should focus not only on better understanding the presentation of mental health conditions in the intellectual disability community and the development and testing of appropriate interventions, but also on ensuring the effective dissemination of findings to the community of practitioners. This can be achieved through improved access to information about research, the creation of research outputs aimed at practitioners, which provide summaries of evidence in non-academic language, and practical advice to translate to practice. For example, see the two-page briefings from the current project aimed at the DXS, FXS and WS communities, and funders, researchers, practitioners and policy makers respectively: https://www.surrey.ac.uk/research-projects/shape-research-change-lives-setting-priorities-genetic-syndrome-research#outputs. Community members’ stated research priorities in terms of service access and interventions is consistent with other priority setting exercises that have worked with intellectual disability and DS communities [13, 14] as well as those on Childhood Disability [15], Learning Difficulties [16] and Childhood Neurological Conditions [17].

#### Researching with the community with lived experience

Our work extended the findings from the existing priority setting-exercises in two important ways. First, we elicited the views and experiences of a range of community members, including parents, professionals, researchers, and most critically, children and adults with DS, FXS and WS themselves. People with genetic syndromes are all-too-often left out of research and of the construction of knowledge about them – as participants, where the perspectives of parents and professionals frequently dominate [12, 14] and as partners in the research process [19]. Ensuring that the perspectives of those with lived experience is captured in priority-setting exercises like this one was therefore crucial to identifying those areas of research that should be prioritised to ensure that they can lead the lives that they wish to lead. Second, our participants did not just identify *what* should be the focus of future research. We also asked about their experiences of research and their views on *how* researchers should seek to conduct their research to address effectively these priorities. Members of the DS, FXS and WS communities reported wanting to see greater involvement of those with lived experience in shaping the research process; greater diversity in those participating in DS, FXS and WS research; research findings that were accessible to the whole community; and better processes to support research collaborations. As noted in the introduction, there is an increasing emphasis on including people with lived experience in research, including basic and more applied science [21, 22, 24]; the current study adds to our knowledge-base of how to increase the relevance and impact of our research.

In Theme 3, parents emphasised the value of research participation which made them feel proactive, gave them better connections to knowledge and experts in the field, as well as information that could be used to access services, which add to existing evidence of the value of participatory approaches to research [21, 22]. However, community members also wanted research to be made more accessible, in a central place that allows them to search for evidence in matters relevant to them. Research driven by lived experience is still in its infancy within genetic syndrome research. Although there was a recognition that there is increasing interest and support from funders for participatory research, the appropriate infrastructure and resources were felt not to be in place. These findings add to the existing literature on inclusive research with the intellectual disability community, which identified challenges in resource allocation, recruitment, and understanding roles and responsibilities [48]. Together, these findings suggest that systemic changes are needed to ensure that participatory research is appropriately accommodated within current academic research frameworks [49]. Furthermore, a deeper understanding of establishing genuine, effective research-community collaborations and sharing power in research is needed in individual researchers and the community members with whom they work.

#### Diversity in genetic syndrome research

Community members, both professionals and parents, highlighted the lack of diversity in genetic syndrome research in Theme 3. They agreed that research often captures the views of a “self-selected” group of individuals, who are not necessarily representative of the voices of the broader community. This adds to Nygaard, Halvorsrud [18]’s criticism that priority-setting exercises often involve individuals associated with charity organisations who are likely to be white, from middle class backgrounds, and with high education attainment levels. Future research should focus on improving access to information about research and on involving those characterised as “difficult to reach”, who are likely to have even greater unmet need and potentially stand to benefit the most from research. In doing so, researchers will need to seek to understand precisely *why* certain groups are seldom heard. Are they hesitant or naïve to what research participation entails? Is the research inaccessible due to digital, geographic, literacy and/or spoken communication barriers? Successful participatory research is founded on strong relationships between academics and community members. Developing trust with seldom-heard groups will take time and sustained effort – and the need for additional resources to do so should be acknowledged by researchers and funders alike. Funding bodies should also work collaboratively with researchers and the communities with lived experience to understand the barriers to conducting meaningful participatory research, and implement changes that support and encourage this type of research.

#### Limitations

Whilst this study was the first to explore the research priorities of the DS, FXS and WS communities and provide valuable insights into how this research should be conducted, it is not without its limitations. Despite efforts from the research team to ensure diversity of perspectives in the present study, the main pathway to recruitment was established by our partner charitable organizations. This meant that most participants had prior experience of taking part in research, and we acknowledge that their views might not be representative of their communities. This lack of diversity is also shown in participant characteristics, with most identifying as white ethnic background and residing in England. Further limitations include the lack of perspectives from children with FXS whom we were not able to recruit for the focus groups, and the data being collected using digital methods and spoken communication. It is possible that the limited sample diversity may have influenced the priorities identified. For example, the under-representation of people with FXS and the absence of children with FXS, may have precluded the possibility of generating syndrome-specific priorities. Similarly, the predominance of English participants from white ethnic backgrounds may have shaped the emphasis on certain forms of service provision, research engagement and access to care. Importantly, the priorities identified in this study reflect the perspectives of the groups represented in the sample and offer meaningful insight into their experiences, while not claiming to capture the full heterogeneity of the DS, FXS, and WS communities. Future priority-setting exercises should seek more diverse samples to ensure that a broader range of voices, backgrounds, and lived experiences are represented.

## Conclusion

To conclude, our priority-setting exercise builds on existing evidence, demonstrating that communities with lived experience in DS, FXS and WS prioritise applied research. That is not to say that they think that basic science research is unimportant. Rather, they simply wanted a more balanced research portfolio, with more research being done that has greater impact on their everyday lives. Our findings also suggest that future efforts should incorporate effective and accessible research translation to facilitate improved public and practitioner awareness and understanding of the unique needs and strengths of individuals. Accessible translations of research findings are not sufficient; outputs for the communities should be easily available in a centralised place such as through the creation of a UK-wide research access hub. Finally, our findings advocate that the value of conducting research “with” the communities needs recognition. This should be accompanied by a shift in understanding DS, FXS and WS research from a conventional medical model framework to a model that acknowledges the strengths and needs of each individual and the contexts in which they are embedded. Researchers should seek to work collaboratively with the communities throughout the research process, and funding bodies should develop infrastructures to support this type of work. Attending to these recommendations should go some way to address the problem identified by Nature [7] – that is, ensuring that research done in the name of DS, FXS and WS is of direct use to these communities – and therefore enabling individuals with these genetic syndromes to lead the lives they deserve to lead.

## Author contributions

LC: Data Curation, Formal Analysis, Investigation, Methodology, Project Administration, Writing – Original Draft Preparation, Writing – review and editing

EP: Conceptualisation, Formal Analysis, Funding Acquisition, Methodology, Supervision, Writing – Original Draft Preparation, Writing – review and editing

JVH: Conceptualisation, Funding Acquisition, Methodology, Writing – review and editing GS: Conceptualisation, Funding Acquisition, Methodology, Writing – review and editing EKF: Conceptualisation, Data Curation, Formal Analysis, Funding Acquisition, Investigation,

Methodology, Project Administration, Supervision, Writing – Original Draft, Writing – review and editing

## Data Availability Statement

We did not gain consent from participants to make the raw qualitative data from this project open, as this may have precluded full discussion of issues that were sensitive to participants. Non-sensitive, non-disclosive data, in the form of anonymised exemplar quotations for all themes/subthemes, is available upon reasonable request from the corresponding authors. If the corresponding authors are unable to respond to requests, interested researchers may contact the University Ethics Committee at the University of Surrey. All relevant study materials are available on OSF at osf.io/db6cg.

## Data Availability

Raw qualitative data cannot be shared publicly because participant consent was not obtained for open-data sharing and to protect the confidentiality of sensitive discussions. Anonymized exemplar quotations supporting the themes and subthemes are available from the corresponding author upon reasonable request. If the corresponding author is unavailable, inquiries can be directed to the University of Surrey Ethics Committee. All relevant study materials are openly accessible on the Open Science Framework at https://osf.io/db6cg

## Footnotes

a Use of language: We use the term “intellectual disabilities” to avoid confusion among the readership, but note that the term “learning disabilities” is preferred in some countries and by some communities, including by some of our own contributors. Our aim is to be precise, but also inclusive and respectful of all perspectives.

b Initially, participants were invited to take part in a group-based discussion as this was seen as the most appropriate method of data collection to help facilitate conversation and present the preliminary findings of the review. Initial expression of interest was minimal from the FXS community, with no participants with FXS willing to take part in the study once provided with further information. A decision was made to offer semi-structured interviews as an alternative, and one adult with FXS took part.

## References

1. Mencap. Mencap’s Big Learning Disability Survey. 2022.

2. Emerson E, Baines S. Health Inequalities & People with Learning Disabilities in the UK, 2010. 2010.

3. Department for Health and Social Care. The UK Rare Diseases Framework. 2021.

4. Abuga JA, Kariuki SM, Kinyanjui SM, Boele van Hensbroek M, Newton CR. Premature Mortality, Risk Factors, and Causes of Death Following Childhood-Onset Neurological Impairments: A Systematic Review. Front Neurol. 2021;12:627824.

5. Emerson E, Hatton C, Baines S, Robertson J. The physical health of British adults with intellectual disability: cross sectional study. International Journal for Equity in Health. 2016;15(1):11.

6. Durbin A, Isaacs B, Mauer-Vakil D, Connelly J, Steer L, Roy S, et al. Intellectual Disability and Homelessness: a Synthesis of the Literature and Discussion of How Supportive Housing Can Support Wellness for People with Intellectual Disability. Current Developmental Disorders Reports. 2018;5(3):125–31.

7. Nature. The best research is produced when researchers and communities work together. Nature. 2018;562(7725):7.

8. Tan DW, Crane L, Haar T, Heyworth M, Poulsen R, Pellicano E. Reporting community involvement in autism research: Findings from the journal Autism. Autism. 2024:13623613241275263.

9. NIHR. UK Standards for Public Involvement. 2019.

10. Crowe S, Fenton M, Hall M, Cowan K, Chalmers I. Patients’, clinicians’ and the research communities’ priorities for treatment research: there is an important mismatch. Research Involvement and Engagement. 2015;1(1):2.

11. Embracing Complexity, Sapiets S. Embracing Complexity in Research on Neurodevelopmental Conditions and Mental Health. 2021.

12. Tomlinson M, Yasamy MT, Emerson E, Officer A, Richler D, Saxena S. Setting global research priorities for developmental disabilities, including intellectual disabilities and autism. J Intellect Disabil Res. 2014;58(12):1121–30.

13. Royal College of Speech and Language Therapists (RCSLT). Learning Disabilities Research Priorities. 2019.

14. Sinclair M, McCullough JE, Elliott D, Latos-Bielenska A, Braz P, Cavero-Carbonell C, et al. Exploring Research Priorities of Parents Who Have Children With Down Syndrome, Cleft Lip With or Without Cleft Palate, Congenital Heart Defects, or Spina Bifida Using ConnectEpeople: A Social Media Coproduction Research Study. J Med Internet Res. 2019;21(11):e15847.

15. Morris C, Simkiss D, Busk M, Morris M, Allard A, Denness J, et al. Setting research priorities to improve the health of children and young people with neurodisability: a British Academy of Childhood Disability-James Lind Alliance Research Priority Setting Partnership. BMJ Open. 2015;5(1):e006233.

16. Lim AK, Rhodes S, Cowan K, O’Hare A. Joint production of research priorities to improve the lives of those with childhood onset conditions that impair learning: the James Lind Alliance Priority Setting Partnership for ‘learning difficulties’. BMJ Open. 2019;9(10):e028780.

17. Cadwgan J, Goodwin J, Babcock B, Brick M, Chin R, Easton A, et al. UK research priority setting for childhood neurological conditions. Developmental Medicine & Child Neurology. 2024;n/a(n/a).

18. Nygaard A, Halvorsrud L, Linnerud S, Grov EK, Bergland A. The James Lind Alliance process approach: scoping review. BMJ Open. 2019;9(8):e027473.

19. Hewitt O, Langdon PE, Tapp K, Larkin M. A systematic review and narrative synthesis of inclusive health and social care research with people with intellectual disabilities: How are co-researchers involved and what are their experiences? Journal of Applied Research in Intellectual Disabilities. 2023;36(4):681–701.

20. Cristescu L, Scerif G, Pellicano L, Van Herwegen J, Farran EK. Shape Research, Change Lives: Setting priorities in genetic syndrome research. 2024.

21. McLaughlin H. Involving young service users as co-researchers: possibilities, benefits and costs. British Journal of Social Work. 2006;36(8):1395–410.

22. Davies S, Staley K. Exploring impact: public involvement in NHS, public health and social care research: Involve; 2009.

23. Oliver K, Kothari A, Mays N. The dark side of coproduction: do the costs outweigh the benefits for health research? Health Research Policy and Systems. 2019;17(1):33.

24. Pellicano E, Lawson W, Hall G, Mahony J, Lilley R, Heyworth M, et al. “I Knew She’d Get It, and Get Me”: Participants’ Perspectives of a Participatory Autism Research Project. Autism in Adulthood. 2021;4(2):120–9.

25. Cristescu L, Farran EK, Van Herwegen J, Scerif G, Pellicano L. Shaping the future of research with individuals with intellectual disability 2023 [Available from: 10.17605/OSF.IO/ZEFKJ.

26. Truscott J, Benton, L. What is a Researcher? [Short Version]: UCL Media Central; 2022.

27. Braun V, Clarke V. Using thematic analysis in psychology. Qualitative research in psychology. 2006;3(2):77–101.

28. Clarke V, Braun V. Successful qualitative research: A practical guide for beginners. 2013.

29. Braun V, Clarke V. Reflecting on reflexive thematic analysis. Qualitative research in sport, exercise and health. 2019;11(4):589–97.

30. McGlinchey E, McCallion P, Burke E, Carroll R, McCarron M. Exploring the issue of employment for adults with an intellectual disability in I reland. Journal of Applied Research in Intellectual Disabilities. 2013;26(4):335–43.

31. Beyer S, Brown T, Akandi R, Rapley M. A comparison of quality of life outcomes for people with intellectual disabilities in supported employment, day services and employment enterprises. Journal of Applied Research in Intellectual Disabilities. 2010;23(3):290–5.

32. Jahoda A, Kemp J, Riddell S, Banks P. Feelings about work: A review of the socio-emotional impact of supported employment on people with intellectual disabilities. Journal of Applied Research in Intellectual Disabilities. 2008;21(1):1–18.

33. McMahon M, Hatton C. A comparison of the prevalence of health problems among adults with and without intellectual disability: A total administrative population study. Journal of Applied Research in Intellectual Disabilities. 2021;34(1):316–25.

34. van Schrojenstein Lantman-de Valk HMJ, Walsh PN. Managing health problems in people with intellectual disabilities. BMJ. 2008;337:a2507.

35. Merrick J, Merrick E. Equal Treatment: Closing the Gap. A Formal Investigation into Physical Health Inequalities Experienced by People with Learning Disabilities and/or Mental Health Problems. Journal of Policy and Practice in Intellectual Disabilities. 2007;4(1):73-.

36. Leonard H, Eastham K, Dark J. Heart and heart-lung transplantation in Down’s syndrome. The lack of supportive evidence means each case must be carefully assessed. 2000;320(7238):816–7.

37. White A, Sheehan R, Ding J, Roberts C, Magill N, Keagan-Bull R, et al. Learning from Lives and Deaths - People with a learning disability and autistic people (LeDeR) report for 2022. The Institute of Psychiatry, Psychology and Neuroscience (IoPPN) King’s College London; 2023.

38. Duff M, Hoghton M, Scheepers M. More training is needed in health care of people with learning disabilities. BMJ. 2000;321(7257):385.

39. Phillips A, Morrison J, Davis RW. General practitioners’ educational needs in intellectual disability health. Journal of Intellectual Disability Research. 2004;48(2):142–9.

40. Macpherson H, Hall E, Power A, Kaley A. Debilitating landscapes of care and support: envisaging alternative futures. Social & Cultural Geography. 2023;24(1):140–56.

41. Pearson C, Ridley J. Is personalization the right plan at the wrong time? Re-thinking cash-for-care in an age of Austerity. Social Policy & Administration. 2017;51(7):1042–59.

42. Cooper S-A, Smiley E, Morrison J, Williamson A, Allan L. Mental ill-health in adults with intellectual disabilities: prevalence and associated factors. British Journal of Psychiatry. 2007;190(1):27–35.

43. Einfeld SL, Ellis LA, Emerson E. Comorbidity of intellectual disability and mental disorder in children and adolescents: A systematic review. Journal of Intellectual and Developmental Disability. 2011;36(2):137–43.

44. Whittle EL, Fisher KR, Reppermund S, Lenroot R, Trollor J. Barriers and enablers to accessing mental health services for people with intellectual disability: A scoping review. Journal of Mental Health Research in Intellectual Disabilities. 2018;11(1):69–102.

45. Costello H, Bouras N. Assessment of mental health problems in people with intellectual disabilities. Israel Journal of Psychiatry and Related Sciences. 2006;43(4):241.

46. Jopp DA, Keys CB. Diagnostic overshadowing reviewed and reconsidered. American Journal on Mental Retardation. 2001;106(5):416–33.

47. Department for Health and Social Care. Reasonable adjustments: a legal duty. 2020.

48. Di Lorito C, Bosco A, Birt L, Hassiotis A. Co-research with adults with intellectual disability: A systematic review. J Appl Res Intellect Disabil. 2018;31(5):669–86.

49. Pickard H, Pellicano E, den Houting J, Crane L. Participatory autism research: Early career and established researchers’ views and experiences. Autism. 2022;26(1):75–87.

